# Large Language Models in Real-World Clinical Workflows: A Systematic Review of Applications and Implementation

**DOI:** 10.1101/2025.06.10.25329323

**Authors:** Yaara Artsi, Vera Sorin, Benjamin S. Glicksberg, Panagiotis Korfiatis, Girish N Nadkarni, Eyal Klang

## Abstract

**Background:** Large language models (LLMs) offer promise for enhancing clinical care by automating documentation, supporting decision-making, and improving communication. However, their integration into real-world healthcare workflows remains limited and under characterized. This systematic review aims to evaluate the literature on real-world implementation of LLMs in clinical workflows, including their use cases, clinical settings, observed outcomes, and challenges.

**Methods:** We searched MEDLINE, Scopus, Web of Science, and Google Scholar for studies published between January 2015 and April 2025 that assessed LLMs in real-world clinical applications. Inclusion criteria were peer-reviewed, full-text studies in English reporting empirical implementation of LLMs in clinical settings. Study quality and risk of bias were assessed using the PROBAST tool.

**Results:** Four studies published between 2024 and 2025 met inclusion criteria. All used generative pre-trained transformers (GPTs). Reported applications included outpatient communication, mental health support, inbox message drafting, and clinical data extraction. LLM deployment was associated with improvements in operational efficiency, user satisfaction, and reduced workload. However, challenges included performance variability across data types, limitations in generalizability, regulatory delays, and lack of post-deployment monitoring.

**Conclusions:** Early evidence suggests that LLMs can enhance clinical workflows, but real-world adoption remains constrained by systemic, technical, and regulatory barriers. To support safe and scalable use, future efforts should prioritize standardized evaluation metrics, multi-site validation, human oversight, and implementation frameworks tailored to clinical settings.

## INTRODUCTION

The integration of large language models (LLMs) into clinical practice has sparked interest across the healthcare community [1]. These technologies have the potential to enhance diagnostic accuracy, reduce administrative burden, and support clinical decision-making [2]. However, while LLMs have demonstrated impressive performance in controlled retrospective settings [3, 4], their translation into clinical workflows remains inconsistent and underexplored [5].

Despite exponential growth, there remains a significant gap between developed models and real-world translation [6]. The majority of explored use cases are still at the proof-of-concept stage, due to regulatory uncertainties, technical deployment barriers, privacy concerns and variable institutional readiness [7, 8].

Moreover, evaluation metrics vary widely across studies, with many reporting model performances in silico without assessing usability, safety, or effectiveness in real-world clinical workflow [9, 10]. There is also a lack of robust post-deployment monitoring systems to better understand the impact and shifting performance of these models.

This systematic review aims to evaluate the existing literature on LLM integration into real-world clinical settings. Specifically, we assess the extent of their deployment, the clinical settings in which they are applied, the tasks they are used for, and the outcomes associated with their use. By doing so, we aim to guide future research and adoption strategies

## METHODS

### Literature Search

We systematically searched the literature to identify studies describing the application of LLMs in a real-world setting. We searched MEDLINE, Google Scholar, Scopus, and the Web of Science for papers published from January 2015 to April 2025. The full search process, including Boolean operators, is detailed in the **Supplementary Materials**.

In addition, we checked the reference lists of selected publications and the “Similar Articles” feature in PubMed, to identify additional publications. Ethical approval was not required, as this is a systematic review of previously published research and does not include individual participant information. Our study followed the Preferred Reporting Items for Systematic Reviews and meta-analyses (PRISMA) guidelines [11]. The study is registered with PROSPERO (CRD420251030069)

### Study Selection

All search results were imported into a single CSV table and deduplicated. Two authors (YA and VS) independently screened titles and abstracts for relevance. Potentially eligible articles were retrieved in full text and assessed by YA and VS. Discrepancies were resolved by a third author (EK).

### Inclusion and Exclusion Criteria

Full-text peer-reviewed publications in English focusing on LLMs integration and deployment in real-world clinical workflow were included. We excluded non-English articles, non-original research, non-peer-reviewed publications, studies that did not assess LLMs, and studies that did not explicitly assess LLMs in real-world settings. **Figure 1** presents the flow diagram of the screening and inclusion process.

**Figure 1.**
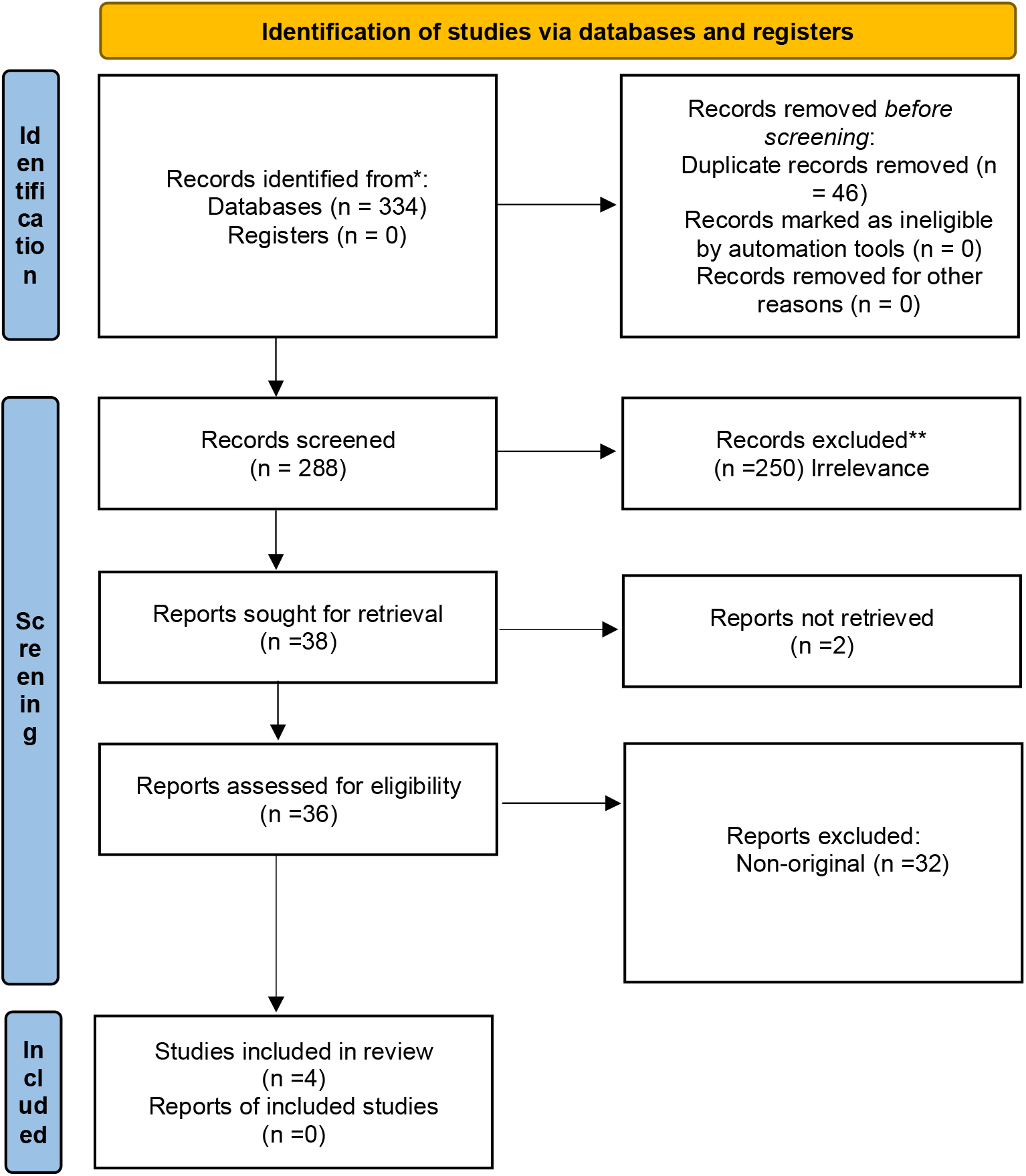
Flow diagram of inclusion and exclusion process

### Quality Assessment

The risk of bias and applicability was evaluated using the PROBAST tool. (**Figure 2 and Figure 3**). A detailed assessment of the studies using the PROBAST tool is detailed in the **Supplementary Material**.

**Figure 2.**
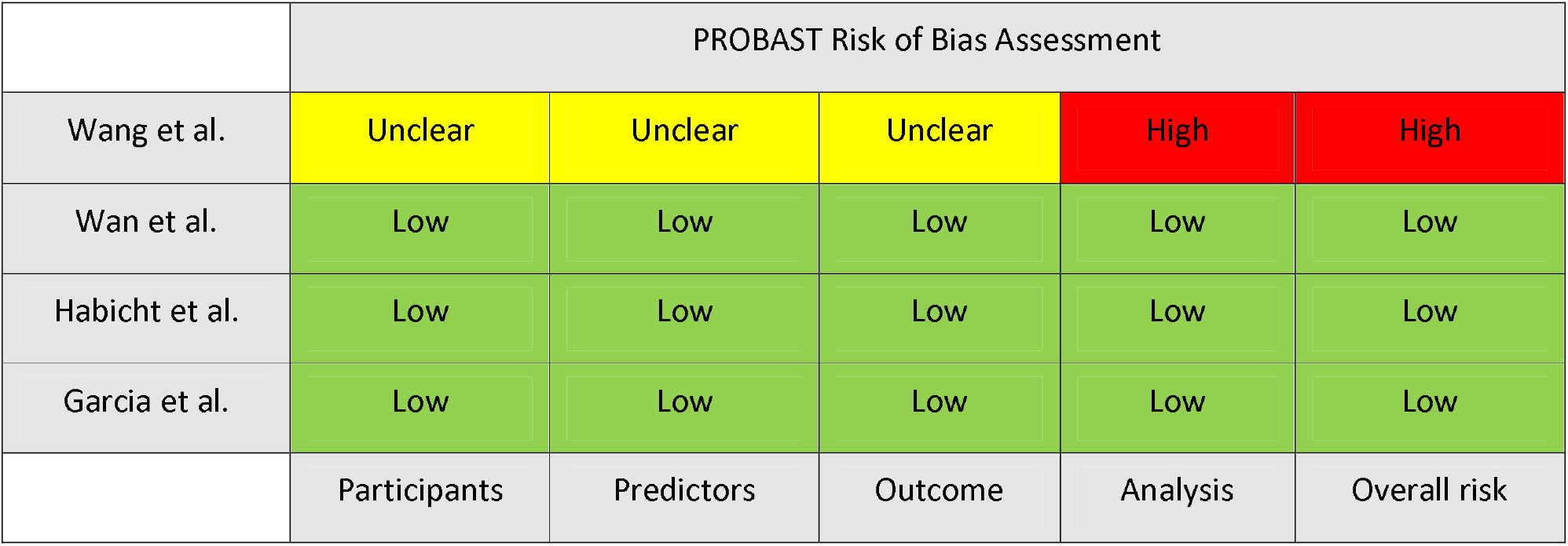
PROBAST Risk of Bias assessment

**Figure 3.**
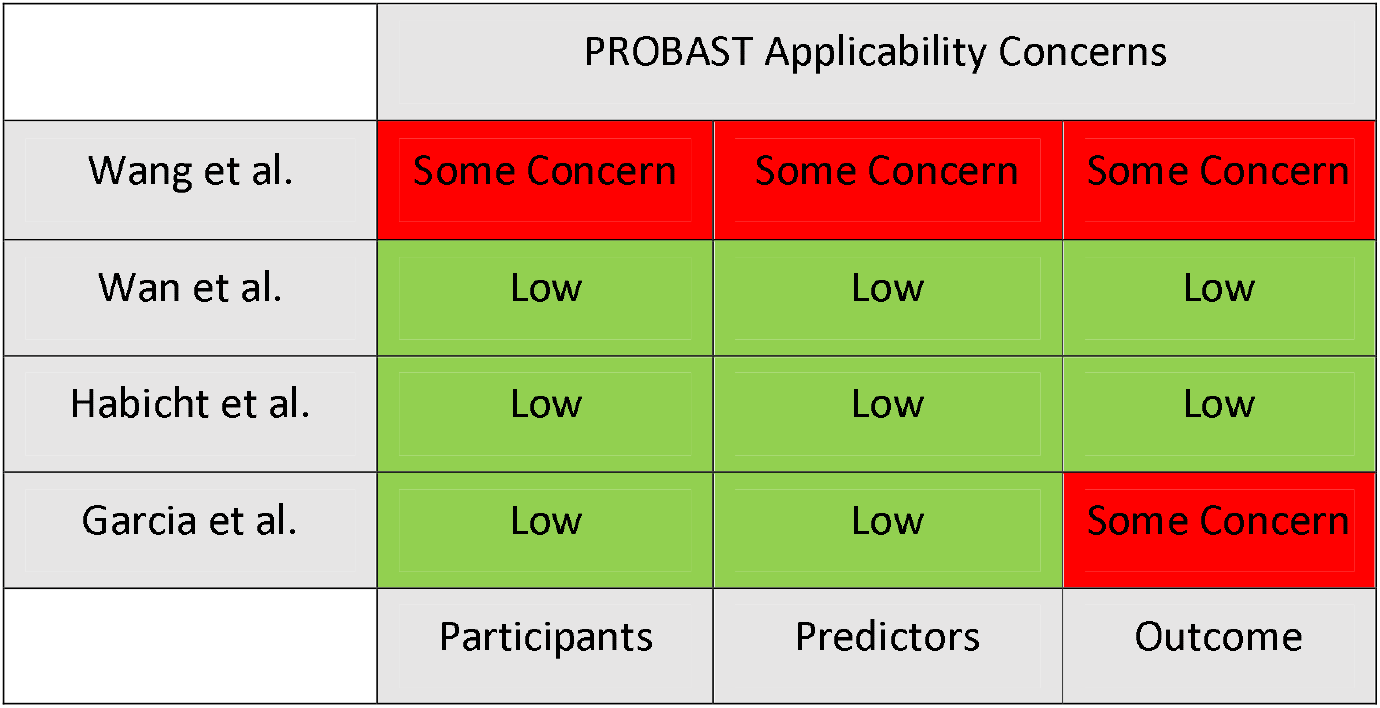
PROBAST Assessment for Applicability

## FUNDAMENTAL CONCEPTS

An overview of fundamental concepts in AI is included in the **Supplementary Material**, along with visual hierarchy shown in **Supplementary Figure 1**.

## RESULTS

### Study Selection and Characteristics

Four studies were included in this review, published between March 2024 and March 2025. All studies utilized generative pre-trained transformers (GPT) (100%). Two studies focused on patient services, including LLM-supported communication during outpatient intake and response to patient messages (50%). Two studies’ focus areas were on data extraction (50%), one applied LLM as a support tool (25%) (**Table 1**). The results of the studies are summarized in **Table 2. Figure 4** provides an overview of the characteristics of the included studies.

**Table 1.** General features of reviewed studies.

| Study | AI Tool | Focus Area | Type of Data | Clinical Setting |
| --- | --- | --- | --- | --- |
| Wang et al<sup>[12]</sup> | ChatGLM2-6B<br>LLaMA2-7B | Data Extraction | Quantitative | Chinese hospital |
| Wan et al<sup>[13]</sup> | Site-Specific Chatbot (SSPEC) | Patient Intake and Reception | Quantitative | Outpatient reception workflows (2 medical centers) |
| Habicht et al<sup>[14]</sup> | GPT-4 | Therapy Support System | Quantitative | Group-based Cognitive Behavioral Therapy (UK Talking Therapies) |
| Garcia et al<sup>[15]</sup> | GPT-3.5<br>Turbo GPT-4 | Clinical Communication Support | Quantitative | Primary care, gastroenterology, hepatology |

**Table 2.**
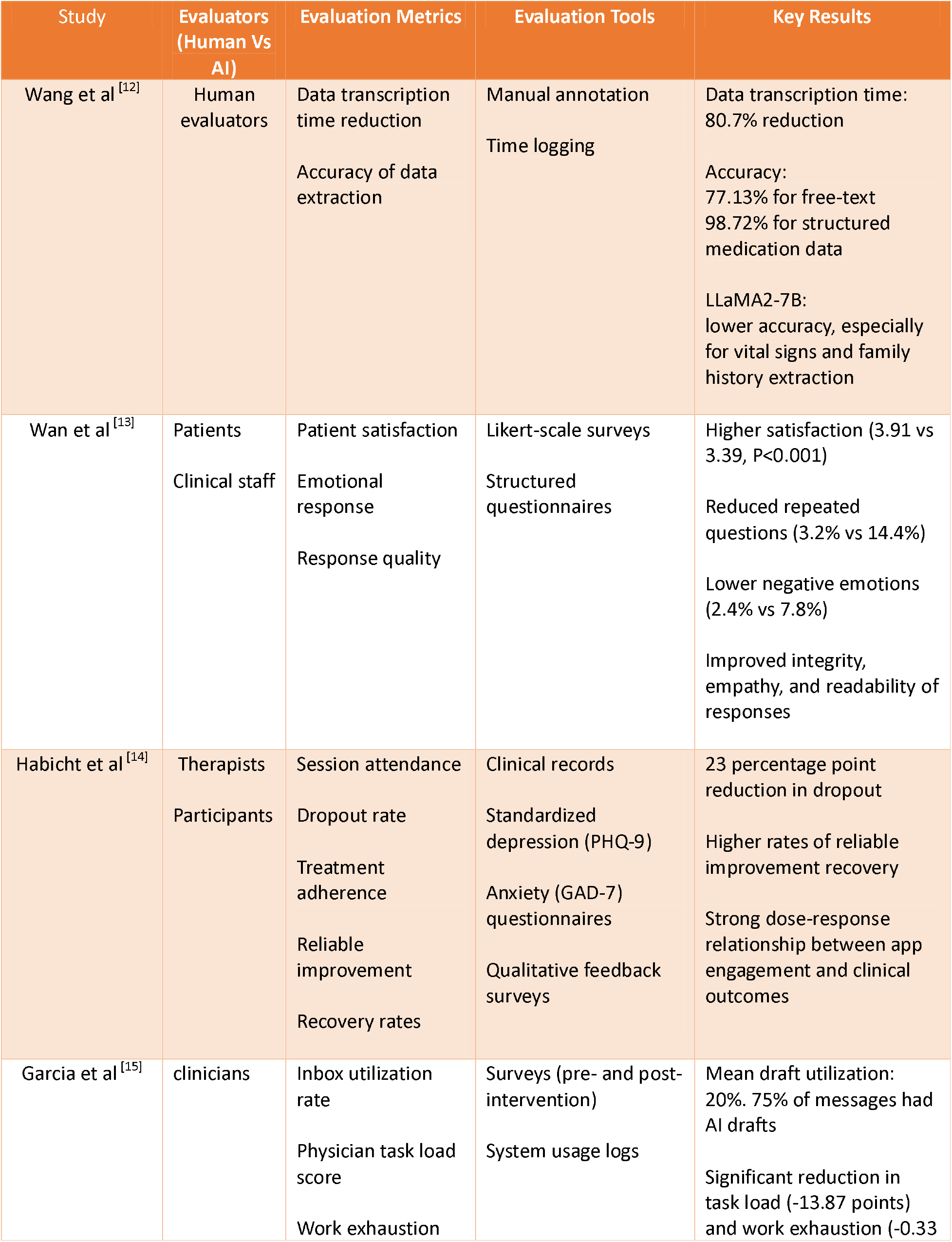
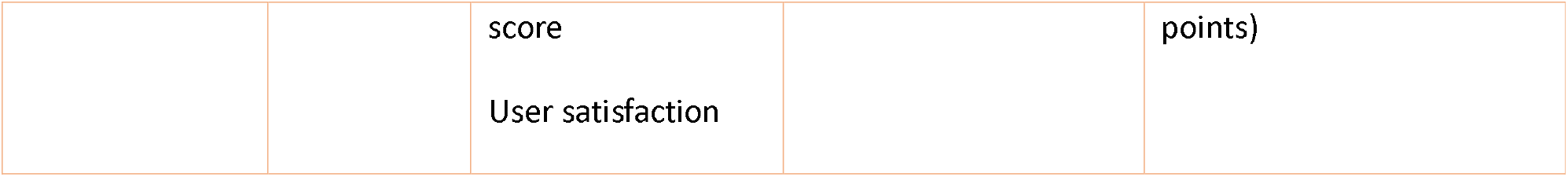
Evaluation Metrics and Key Results of Reviewed Studies.

| Study | Evaluators<br>(Human Vs<br>AI) | Evaluation Metrics | Evaluation Tools | Key Results |
| --- | --- | --- | --- | --- |
| Wang et al<sup>[12]</sup> | Human<br>evaluators | Data transcription<br>time reduction<br>Accuracy of data<br>extraction | Manual annotation<br>Time logging | Data transcription time:<br>80.7% reduction<br>Accuracy:<br>77.13% for free-text<br>98.72% for structured<br>medication data<br>LLaMA2-7B:<br>lower accuracy, especially<br>for vital signs and family<br>history extraction |
| Wan et al<sup>[13]</sup> | Patients<br>Clinical staff | Patient satisfaction<br>Emotional<br>response<br>Response quality | Likert-scale surveys<br>Structured<br>questionnaires | Higher satisfaction (3.91 vs<br>3.39, P<0.001)<br>Reduced repeated<br>questions (3.2% vs 14.4%)<br>Lower negative emotions<br>(2.4% vs 7.8%)<br>Improved integrity,<br>empathy, and readability of<br>responses |
| Habicht et al<sup>[14]</sup> | Therapists<br>Participants | Session attendance<br>Dropout rate<br>Treatment<br>adherence<br>Reliable<br>improvement<br>Recovery rates | Clinical records<br>Standardized<br>depression (PHQ-9)<br>Anxiety (GAD-7)<br>questionnaires<br>Qualitative feedback<br>surveys | 23 percentage point<br>reduction in dropout<br>Higher rates of reliable<br>improvement recovery<br>Strong dose-response<br>relationship between app<br>engagement and clinical<br>outcomes |
| Garcia et al<sup>[15]</sup> | clinicians | Inbox utilization<br>rate<br>Physician task load<br>score<br>Work exhaustion | Surveys (pre- and post-<br>intervention)<br>System usage logs | Mean draft utilization:<br>20%. 75% of messages had<br>AI drafts<br>Significant reduction in<br>task load (-13.87 points)<br>and work exhaustion (-0.33 |
|  |  | score |  | points) |
|  |  | User satisfaction |  |  |

**Table 3.** Limitations and Challenges in Reviewed Studies.

| Study | Challenges | Domain Impact | Observed or Inferred |
| --- | --- | --- | --- |
| Wang et al<sup>[12]</sup> | Moderate accuracy variability (77.13% for free-text extraction)<br>Lower performance compared to LLaMA2-7B in some fields | Data extraction accuracy | Observed |
| Wan et al<sup>[13]</sup> | Need for careful prompt design<br>Reliance on nurse oversight | Patient Communication<br>Workflow Efficiency<br>Scalability and generalizability | Inferred |
| Habicht et al<sup>[14]</sup> | App retention decline (only 19.3% engaged by week 6) | Mental Health Outcomes<br>Patient Engagement<br>Tool sustainability | Observed |
| Garcia et al<sup>[15]</sup> | Variability in tone, message relevance, and adoption<br>Modest impact on inbox time<br>Concerns about over-reliance on AI | Administrative Burden<br>Clinician Well-being | Observed |

**Figure 4.**
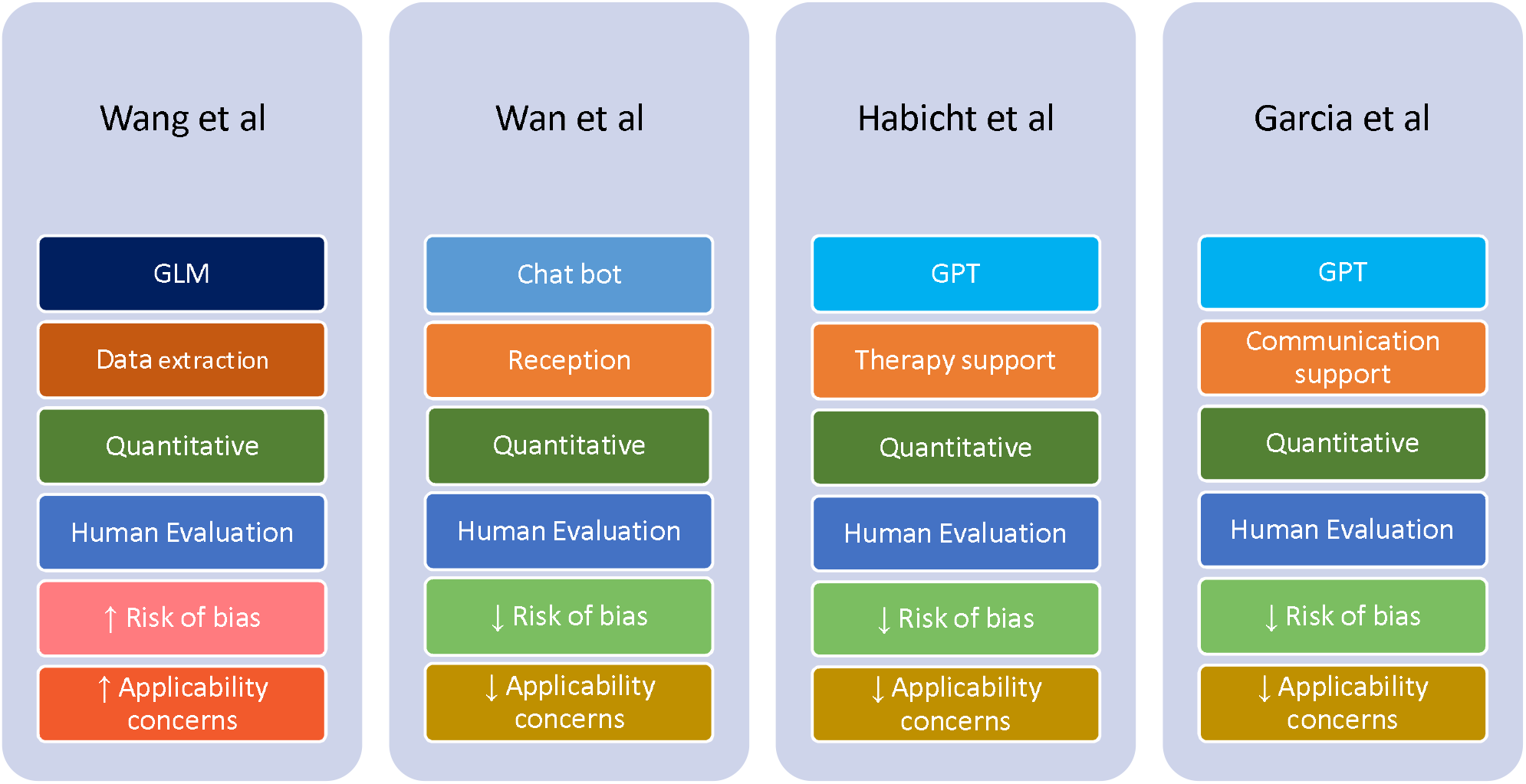
Differences and Similarities in Various Parameters Presented in the Reviewed Studies

### Descriptive summary of results

Wang et al [12] evaluated ChatGLM2-6B for real-world data extraction. The model achieved an 80.7% reduction in transcription time. The accuracy varied by data type, 77.13% for free text and 98.72% for structured medication data. In comparison, the LLaMA2-7B model showed lower accuracy, especially for vital signs and family history (**Table 2**).

Wan et al [13] assessed a site-specific chatbot (SSPEC) integrated into outpatient workflows across two medical centers, comparing it to traditional nurse-only interactions. The nurse–SSPEC model improved patient satisfaction, reduced repeated questions (3.2% vs. 14.4%), and lowered negative patient emotions (2.4% vs. 7.8%). It also enhanced response quality in integrity, empathy, and readability (**Table 2**).

Habicht et al [14] assessed a GPT-4–powered AI tool in group-based CBT and found it improved clinical outcomes. The AI group had more session attendance, fewer missed appointments, and a 23-percentage point lower dropout rate than those using standard worksheets—higher engagement correlated with better adherence and outcomes. Qualitative feedback also noted improved self-awareness, mindfulness, and practical use of CBT techniques (**Table 2**).

Garcia et al [15] evaluated GPT-3.5 Turbo and GPT-4 for generating draft replies to patient messages in gastroenterology, hepatology, and primary care. The AI drafts improved efficiency and reduced clinician workload without compromising communication quality. Among 162 clinicians, draft utilization averaged 20%, with 75% of messages receiving AI-generated replies. While time spent on inbox tasks did not significantly change, clinicians reported reduced task load and work exhaustion. User feedback raised concerns about message tone, length, and relevance (**Table 2**).

## DISCUSSION

LLMs show promise in real-world clinical workflows [16], with the potential to enhance many fields in clinical care (**Figure 5**). However, their implementation remains early-stage and context-dependent. Most of the studies we reviewed reported positive outcomes, yet critical challenges that affect generalizability, integration, and long-term sustainability remain.

**Figure 5.**
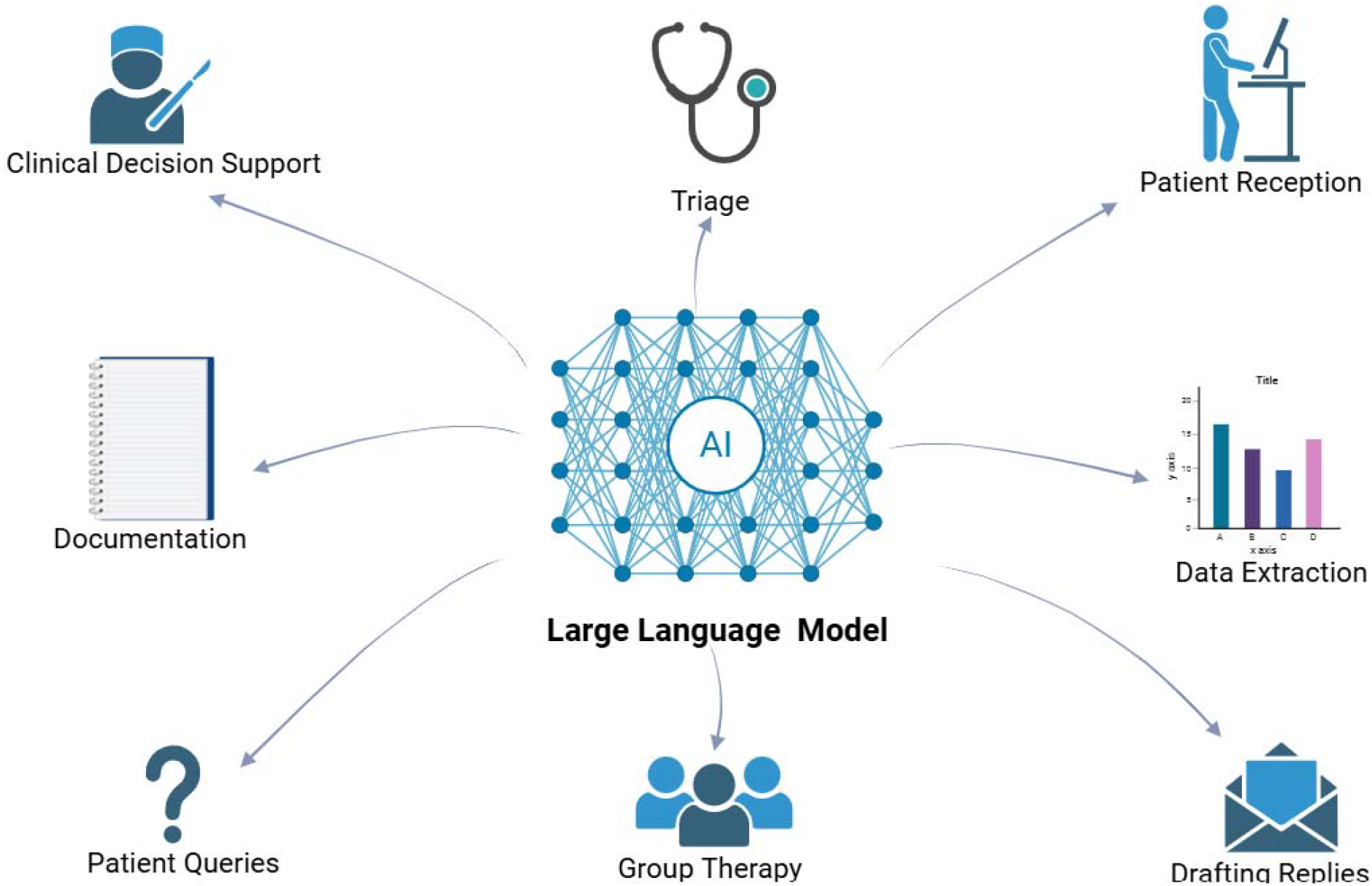
LLM application in various fields in real-world setting

Several studies demonstrated clear empirical benefits. For example, Habicht et al. [14] showed that GPT-4 as a therapy support tool reduced therapy dropout rates and improved clinical outcomes. Wan et al. [13] reported that the site-specific LLM chatbot (SSPEC) reduced repeated interactions and negative patient emotions. Garcia et al. [15] found that GPT-generated draft replies decreased clinician task load and work exhaustion across several clinical settings.

Despite these successes, limitations were reported as well. In Wang et al. [12], while ChatGLM2-6B achieved a reduction in transcription time, accuracy varied significantly across data types. LLaMA2-7B performed notably worse in comparison. These discrepancies accentuate the importance of task specificity and local calibration in AI deployment.

While LLMs can improve operational efficiency and clinical outcomes, widespread adoption is limited by systemic barriers and arbitrary evaluation metrics [17]. These include heterogeneous EHR systems and unclear standards for performance evaluation. Future research should focus on efficient prospective validation and developing clear standardized evaluation metrics. We propose a list of possible evaluation metrics detailed in **Table 4**.

**Table 4.** Suggestions for Standardize Evaluation Metrics.

| Domain | Recommended Metric |
| --- | --- |
| Model performance | AUROC<br>Accuracy<br>Precision<br>Recall<br>F1-score |
| Clinical impact | Patient outcomes |
| Workflow efficiency | Task time reduction<br>Response delay |
| Useability | Clinician reported task load |
| Reliability | Rate of false negatives/positives<br>Override rates |
| Deployment fidelity | User adherence rates<br>Percentage of AI-assisted tasks |
| Generalizability | Cross-site performance variance<br>Calibration curves |

Translating LLMs from experimental settings into clinical practice remains a challenge. Clinical implementation is frequently delayed by regulatory barriers, including classification as software-as-a-medical-device (SaMD), which necessitates lengthy regulatory approval processes and extensive local validation [18]. These processes contribute to version lag, whereby newer models become available before prior versions are deployed or evaluated. Furthermore, performance may degrade over time due to evolving clinical documentation, user behavior, or patient populations [19]. These challenges underscore the need for post-deployment monitoring frameworks and standardized scientific reporting to ensure model safety, performance, and generalizability in clinical environments [20,21].

### Strategies for the future

Several strategic approaches should be considered to facilitate the broader adoption of LLM technologies in clinical workflows. Local adaptation of models is essential, as performance can vary significantly depending on institutional characteristics such as data quality, documentation styles, and patient demographics. Ensuring that models are trained and validated on local data can improve generalizability and clinical relevance. Also, incorporating a human expert oversight framework can improve safety, accountability, and user trust.

AI tools should be designed with task specificity; models tailored to distinct clinical functions, such as triage, documentation, or medication extraction, are more likely to achieve meaningful utility. This can be achieved using fine-tuning [22] or Retriever-Augmented Generation (RAG) [23].

A standardized set of metrics should be developed to support consistency and facilitate future evaluations. We propose our set of metrics in **Table 4**. Also, involving clinicians and end users in the development and implementation process ensures that tools are aligned with real-world workflow needs, increasing user acceptance. Collecting user feedback during deployment can guide iterative model refinement and usability improvements.

Safety mechanisms and override options are crucial to prevent unintended consequences such as clinical errors, automation over-reliance, data bias, and alert fatigue [24]. These risks can disrupt workflows and raise ethical concerns about accountability. Human oversight and ongoing monitoring are essential to ensure AI supports, rather than compromises, clinical care [25].

Ensuring that AI tools are clinically effective requires more than technical performance. This includes interoperability with EHRs, standardized evaluation metrics, and human-centered design features like transparency and clinician oversight. By applying these strategies, AI can move beyond experimental use to become a trusted part of routine clinical care

This review has several limitations. First, the number of eligible studies evaluating LLMs in real-world clinical workflows remains limited, reflecting the early stage of implementation research in this domain. Second, the heterogeneity in study design, evaluation methods, clinical settings, and outcome reporting precluded formal meta-analysis. Third, most included studies were conducted in high-resource settings, potentially limiting the generalizability of findings to low- and middle-income countries. Additionally, some of the studies relied on self-reported outcomes or lacked long-term follow-up, which may introduce reporting bias or fail to capture sustained clinical impact. Finally, despite efforts to capture a comprehensive set of studies, some relevant work may have been missed due to language restrictions, database coverage, or publication lag.

## Conclusion

LLMs demonstrate early but uneven success in real-world integration, with empirical improvements in efficiency and user satisfaction. However, challenges related to generalizability, interoperability, and evaluation must be addressed to ensure scalable and safe adoption. Future research should prioritize multi-center validation, standardized metrics, and end-user collaboration to support the responsible and effective use of AI in clinical care.

## Data Availability

All data produced in the present work are contained in the manuscript

## Abbreviations

AI: Artificial Intelligence
CBT: Cognitive Behavioral Therapy
EHR: Electronic Health Record
GPT: Generative Pre-trained Transformer
LLM: Large Language Model
NHS: National Health Service
NLP: Natural Language Processing
PHQ-9: Patient Health Questionnaire-9
PROBAST: Prediction model Risk of Bias Assessment Tool
SSPEC: Site-Specific Prompt Engineering Chatbot

## Legends

Figure 1. Depict inclusion and exclusion process of reviewed studies

Figure 2. Risk of bias using the PROBAST tool

Figure 3. Applicability concerns using the PROBAST tool

Table 1. Summary of reviewed studies and relevant characteristics

## Declarations

### Ethics approval and consent to participate

Not applicable

### Consent for publication

Not applicable.

### Availability of data and materials

All data generated or analyzed during this study are included in this published article and its supplementary information files.

### Competing interests

The authors declare that they have no competing interests

### Funding

Not applicable

### Authors’ contributions

YA conducted the literature search, data synthesis and wrote the primary manuscript. VS, BSG, and PK reviewed and edited the manuscript. GNN and EK contributed to the conceptualization, data synthesis, and editing of the manuscript. All authors read and approved the final man uscript.

## Notes

### Competing Interest Statement

The authors have declared no competing interest.

### Funding Statement

This study did not receive any funding

