## Supplementary Material for "Large Language Models in Real-World Clinical Workflows: A Systematic Review of Applications and Implementation"

### Fundamental Concepts Overview

Artificial Intelligence (AI) encompasses any technique that allows machines to simulate human intelligence through rules, logic, statistics, or learning [1]. Within AI, Machine Learning (ML) refers to a subset that focuses specifically on systems capable of learning from data to improve performance over time [2].

A further subfield of ML is deep learning, which uses multi-layered neural networks to model complex patterns. Natural language processing (NLP) is an application area within AI and ML that deals with understanding and generating human language [3]. Large Language Models (LLMs) are specific types of deep learning models trained on vast amounts of text data to perform various NLP tasks [4].

#### Supplemental Figure 1 Visual hierarchy of categories in AI

AI

Natural Language Processing

Machine Learning

Deep Learning

Large Language Model

### Literature search process

The search was conducted using the following Boolean operators:

("large language model" OR "large language models" OR ChatGPT OR "GPT-4" OR "GPT-3" OR BERT OR "transformer model" OR "foundation model")

AND

("real-world evidence" OR "real world application" OR "clinical implementation" OR "routine practice" OR "clinical use" OR deployment OR "workflow integration")

AND

("clinical practice" OR "healthcare setting" OR hospital OR "medical setting")

AND

("original research" OR "observational study" OR "clinical study" OR "implementation study")

### PROBAST detailed assessment

#### Wang et al

1. Participants

The study involves real-world clinical data from Chinese hospitals using Electronic Source Data Repository (ESDR) systems, with a focus on evaluating ChatGLM-assisted data extraction. However, the criteria for selecting the medical records or forms (free-text vs. structured) used in the evaluation are not explicitly defined. This makes it difficult to assess potential selection bias.

Risk of bias: **Unclear**

2. Predictors

The LLM (ChatGLM) was used to extract specific clinical data from forms, including free-text entries and structured discharge medication data. The predictors (i.e., textual data inputs) are relevant to the clinical task. However, the study lacks information on how consistently these predictors were formatted or annotated, and whether human input was standardized.

Risk of bias: **Unclear**

3. Outcome

The outcomes measured include extraction accuracy and reduction in transcription time. These outcomes are directly relevant to the tool's performance, but outcome definitions (e.g., criteria for accuracy, who verified correctness) are not fully described.

Risk of bias: **Unclear**

4. Analysis

The study provides basic accuracy metrics and compares ChatGLM performance with LLaMA2-7B, noting that ChatGLM outperformed LLaMA in most tasks. However, statistical validation methods, error analysis, and confidence intervals are not provided. No cross-validation or external validation is discussed, which raises concerns about analytical rigor.

Risk of bias: **High**

Overall Assessment

Risk of Bias: Due to limited methodological detail in the analysis and unclear bias in other domains. Overall risk of bias is assessed as **high**.

Applicability: While the study reflects real-world clinical data extraction settings, it is specific to Chinese hospital infrastructure and may not generalize to other countries or EHR systems. Therefore, applicability is **moderate**.

#### Wan et al

1. Participants

The study involved 2,164 outpatients across two medical centers in China, randomized into a nurse-only group and a nurse–LLM collaboration group (SSPEC). Inclusion criteria were individuals attending outpatient reception services; exclusion criteria were not extensively detailed. While randomization minimizes selection bias, limited detail on baseline characteristics and exclusions introduces slight uncertainty.

Risk of bias: **Low to unclear**

2. Predictors

The predictor in this context was the SSPEC LLM-generated response integrated into the outpatient workflow. The study does not clearly explain the consistency of prompts used or the standardization of LLM outputs across sites. However, the deployment was embedded in a structured clinical workflow, which supports reliability.

Risk of bias: **Unclear**

3. Outcome

Outcomes were clearly defined and measured: patient satisfaction, repeated Q&A rates, emotional expression, and quality of nurse-patient communication (assessed via integrity, empathy, and readability ratings). These were quantitatively assessed using validated metrics, with statistically significant differences between groups.

Risk of bias: **Low**

4. Analysis

The analysis included appropriate statistical comparisons with reported p-values and confidence intervals. However, the study lacks external validation or replication in other settings, and while performance metrics are robust, more detail on the statistical models (e.g., adjustment for confounders) would enhance interpretability.

Risk of bias: **Low to moderate**

Overall Assessment

Risk of Bias: The study is well-conducted, with outcome measures and clear randomization, though more transparency in predictor consistency and generalizability would strengthen confidence. Overall risk of bias is **low to moderate**.

Applicability: The study tests LLM use in real outpatient clinical workflows, directly aligning with real-world implementation questions. Generalizability beyond the two hospitals may be limited by cultural or linguistic factors in SSPEC’s development. Overall applicability is **high**.

#### Habicht et al

1. Participants

The study included 244 patients undergoing group-based cognitive behavioral therapy (CBT) in the UK's NHS Talking Therapies. Participants self-selected into either the intervention group (AI-enabled support tool) or the control group (traditional static materials). Although demographic data and baseline scores were compared and statistically controlled, self-selection may introduce bias due to non-random group assignment.

Risk of bias: **High**

2. Predictors

The predictor was the use of the GPT-4–powered Limbic Care app. While the tool was standardized across participants and tracked usage metrics (e.g., app retention, engagement with exercises), there is limited detail on prompt consistency or variability in patient interactions.

Risk of bias: **Low to unclear**

3. Outcome

Outcomes included treatment adherence (e.g., number of sessions attended, dropout rate) and clinical improvement (e.g., PHQ-9, GAD-7 scores). These were objectively measured and statistically compared, with appropriate controls for baseline scores.

Risk of bias: **Low**

4. Analysis

The analysis applied appropriate statistical tests, reported confidence intervals, and adjusted for confounders (e.g., gender, sexuality, baseline depression/anxiety). A dose-response relationship was explored. However, the observational nature of the study and lack of randomization limit causal inference.

Risk of bias: **Moderate**

Overall Assessment

Risk of Bias: Due to non-randomized group assignment and potential self-selection bias, though outcome measurement and analysis were strong. Overall risk of bias is **moderate to high**.

Applicability: The study was conducted within a real NHS setting with actual patients, supporting relevance for real-world implementation of LLMs in therapy contexts. Applicability is **high**.

#### Gracia et al

1. Participants

The study enrolled 197 ambulatory care clinicians across gastroenterology, hepatology, and primary care settings. Of these, 162 were included in the final analysis. Inclusion criteria were well-documented and based on role and engagement in clinical messaging workflows. Exclusions (e.g., out-of-office status, triage roles) were clearly justified. The participant selection is appropriate for evaluating clinician-facing AI tools.

Risk of bias: **Low**

2. Predictors

The predictor was exposure to GPT-4–generated draft replies integrated into the EHR message system. Usage was automatically logged, and subgroup utilization was analyzed. The study clearly defines the nature and deployment of the intervention.

Risk of bias: **Low**

3. Outcome

Outcomes included clinician time spent in the inbox, message utilization rates, perceived workload (task load score), and burnout (work exhaustion score). Both objective (EHR log data) and subjective (surveys) measures were used. Pre-post comparisons were made, and appropriate scales were applied.

Risk of bias: **Low**

4. Analysis

The study employed descriptive statistics, Kruskal-Wallis tests, and linear mixed effects models to evaluate pre-post differences and subgroup variability. While results were statistically significant for key outcomes (e.g., workload reduction), the observational design limits causal inference. Nevertheless, the methodology is appropriate for implementation evaluation.

Risk of bias: **Low to moderate**

Overall Assessment

Risk of Bias: The study design clearly defined intervention, outcomes, and valid statistical approach. Overall risk of bias is **low**.

Applicability: The study was conducted in real ambulatory care settings using a production EHR system, with outcomes relevant to clinician workflow and well-being. The applicability is **high**
